# The Evolution and Equity of China’s Pharmacist Workforce in Healthcare Institutions: A Provincial Panel Data Analysis, 2007–2023

**DOI:** 10.64898/2026.04.22.26351514

**Authors:** Yuxiang Xia, Lulu Sun, Yingbo Zhao

**Affiliations:** CountryDepartment of Pharmaceutical Administration Research,National Institute of Hospital Administration,NHC,National Cen-ter for Medical Quality Control in Pharmaceutical Management,Beijing, China

**Keywords:** China, Pharmacist, Health human resources, Fairness assessment, Moran’s I

## Abstract

**Background:** China has implemented policies to strengthen its pharmacist workforce since the 2009 healthcare reform, yet a comprehensive evaluation of their long-term systemic effects is lacking.

**Objective:** To systematically analyze the evolution of China’s pharmacist workforce in healthcare institutions from 2007 to 2023 across four dimensions: quantity, quality, structure, and distribution, providing an empirical foundation for policy optimization.

**Methods:** A retrospective analysis was conducted using longitudinal data from the China Health Statistics Yearbooks. Trends were delineated via descriptive statistics. Equity and spatial evolution were assessed using the Gini coefficient, Theil index decomposition, and spatial autocorrelation analyses (Global Moran’s I and hotspot analysis).

**Results:** From 2007 to 2023, the total number of pharmacists increased from 357,700 to 569,500 (average annual growth: 2.2%). This growth lagged behind physicians (4.6%) and nurses (7.4%),causing the pharmacist-to-physician ratio to decline from 1:5.15 to 1:8.39. The workforce showed trends of feminization (female proportion rose from 59.7% to 70.8%) and aging. While quality improved, 51.1% still held an associate degree or below, and only 6.6% held senior titles. Equity analysis revealed the provincial Gini coefficient improved from 0.145 to 0.093. Theil index decomposition confirmed intra-provincial disparities as the primary inequality driver. Spatial analysis showed a non-significant global Moran’s I by 2023 (0.154, *P*\*>0.05), down from 0.254 (*P*<0.01) in 2007. Hotspot analysis confirmed this transition, revealing a contraction of high-confidence clusters and a trend toward balanced distribution.

**Conclusions:** China has made measurable progress in expanding pharmacist workforce size and improving inter-provincial equity since 2007. However, persistent structural challenges remain: relative workforce contraction compared to other health professions, an aging demographic, a shortage of senior talent, and significant intra-provincial inequity. Future policies must prioritize optimizing workforce structure and enhancing clinical service capabilities to catalyze a shift toward patient-centered pharmaceutical care.

**Highlights:**

1. First longitudinal study (2002–2023) tracking China’s institutional pharmacist workforce post-healthcare reform, revealing a critical structural shortage.
2. Pharmacist growth rate (2.2% annually) severely lagged physicians (4.6%) and nurses (7.4%), causing the pharmacist-to-physician ratio to plummet from 1:5.15 to 1:8.39.
3. 69.2% of China’s drug market (prescription drugs) is managed by only 569,500 institutional pharmacists—175,000 fewer than retail pharmacists, exposing a critical workload imbalance.
4. Spatial disparity paradox: Gini coefficient improved to 0.093 (high equity), yet Theil decomposition revealed intra-provincial (urban/rural) gaps as the primary driver of inequality.
5. High-level talent deficit: Despite quality gains, only 6.6% hold senior titles and 6.1% have master’s degrees—a bottleneck for advancing clinical pharmaceutical care.

## 1 Introduction

The Global Burden of Disease Study (GBD) published in The Lancet identifies chronic non-communicable diseases (NCDs) as the leading threat to global health, the effective control of which relies heavily on long-term, standardized, and safe pharmacotherapy [1]. Within this context, pharmacists, as core professionals in medication therapy management, are poised to play a pivotal role in ensuring safe and effective medication use, improving patient outcomes, and supporting healthcare systems in meeting escalating service demands. The World Health Organization (WHO) and the International Pharmaceutical Federation (FIP) have outlined a model for hospital pharmacists, conceptualized as the “Eight-Star Pharmacist,” which encompasses the roles of care provider, communicator, decision-maker, manager, leader, researcher, educator, and lifelong learner [2]. This framework mandates that pharmacists assume greater responsibility for medication-use outcomes and continuously develop their capacity to deliver expanded patient-centered services, with the ultimate goal of achieving safe, effective, and economical medication use. This model defines the requisite competencies and sets the direction for the future development of the pharmacy profession.

Unlike the unified pharmacist management system implemented in most countries, China operates a distinctive “dual-track” system. This system comprises two parallel frameworks: a professional and technical title qualification system for pharmacists overseen by the health administration, and a licensed pharmacist qualification system regulated by the drug regulatory authorities.

Pharmacists working within healthcare institutions fall under the jurisdiction of the national health administrative system regarding their qualification certification, professional titles, and practice management. They are integral members of the clinical care team, with a core mandate to ensure rational medication use. Conversely, “Licensed Pharmacists” registered in community retail pharmacies are managed by the drug regulatory authorities. They primarily practice at the retail end of the pharmaceutical supply chain, with their professional focus on public-facing prescription dispensing, medication counseling, and pharmaceutical care.

This study concentrates specifically on the pharmacist workforce within healthcare institutions. This focus is justified on two principal grounds: first, the consistency and comparability of data sources, as all data are derived from the national health statistics system; second, within the context of deepening healthcare reform, institutional pharmacists are pivotal to undertaking the transformation of pharmaceutical care, participating in multidisciplinary treatment, and influencing patient outcomes. Their allocation and development directly bear on the quality and safety of medical care.

To meet the escalating demands for healthcare, China has established the world’s largest healthcare service system, disease prevention and control network, and healthcare insurance scheme since the launch of the comprehensive healthcare system reform in 2009 [3-4]. Throughout this process, the development of the pharmacist workforce has received sustained attention, with its growth consistently steered by national policy guidance.

In 2018, the National Health Commission issued the Opinions on Accelerating the High-Quality Development of Pharmaceutical Care, which advocated for a strategic shift in pharmaceutical care from being “drug-centered” to becoming “patient-centered,” explicitly emphasizing the need to strengthen the pharmacist workforce [5]. Subsequently, the *14th Five-Year Plan for Health Human Resources Development*, released in 2022, formally integrated the pharmacist workforce into the national blueprint for health human resources. It set a clear quantitative target: through enhanced allocation and training efforts, the total number of pharmacists in healthcare institutions should reach 770,000 by 2025 [6].

Driven by this series of policies, China’s pharmacist workforce has continued to expand. According to the Statistical Bulletin on the Development of Health Care in China 2025, healthcare institutions nationwide employed a total of 592,000 pharmacists by the end of 2024 [7]. Compared to the target set for the end of the “14th Five-Year Plan”period, a discernible gap remains, underscoring the necessity for continued and strengthened workforce development efforts.

However, the inequitable distribution of the health workforce is a persistent global crisis. In China, this issue is exacerbated by its vast population, extensive geographical area, and rapidly accelerating aging demographic [8-11].

Longitudinal studies indicate that despite an overall improvement in the allocation of health technicians in Chinese hospitals and primary care institutions between 2010 and 2021, the per capita availability in the western region exceeded that of the eastern and central regions. Furthermore, the equity of distribution based on geographical area was substantially lower than that based on population distribution [12].

Spatial analyses further reveal pronounced agglomeration characteristics in the distribution of both physician and nurse resources in China. Specifically, physician resources exhibit a polarized pattern with high-value clusters in northern regions and low-value clusters in the south. Similarly, nurse resources demonstrate a dense concentration in northern areas, particularly in Beijing [13]. Although China’s nursing workforce achieved significant progress in both scale and quality from 1998 to 2018, persistent challenges remain, including the attrition of young nurses and marked regional disparities in distribution [14].

While these studies have provided in-depth analyses of the distribution of physician and nursing human resources, dedicated investigations into the critical group of pharmacists remain notably insufficient. The limited existing research on pharmacists suffers from several distinct shortcomings. Temporally, most studies fail to cover the complete cycle since the 2009 healthcare reform, thus limiting the assessment of long-term policy effects. Analytically, they predominantly focus on quantitative distribution, paying insufficient attention to deeper, qualitative transformations within the workforce—such as changes in quality structure and the evolution of service models. This overlooks the policy-driven transition of pharmacists’ roles from primarily “drug dispensing” to “clinical pharmaceutical care,” as advocated under the national initiative for high-quality development of pharmaceutical care. Furthermore, from a conceptual perspective, there is a lack of mechanistic exploration that explicitly links the evolution of the pharmacist workforce to the trajectory of national macro-level policy processes [15-18].

Given the overarching goal of China’s new healthcare reform guidelines and plans to promote a more equitable and efficient allocation of medical resources, investigating disparities and inequities in resource distribution since the 2009 reform is of critical importance, especially as the reform enters a pivotal phase.

This study conducts an in-depth analysis of the changes in the quantity, quality, structure, and distribution of the pharmacist workforce within Chinese healthcare institutions from 2007 to 2023. It examines the impact of relevant policies and uncovers underlying issues, thereby providing an empirical basis for optimizing the allocation of healthcare human resources and promoting regional equity.

## 2 Methods

### 2.1 Health workforce definition

In this study, “pharmacist” refers to all qualified pharmaceutical professionals practicing within healthcare institutions in China. This definition strictly follows the official Chinese standard Health Statistical Indicators (WS/T 598.9-2018) to ensure data consistency. The standard formally defines this group as “药师 (士)” — meaning professionals holding appointed titles such as Assistant Pharmacist, Pharmacist, Pharmacist-in-Charge, Associate Chief Pharmacist, or Chief Pharmacist, while excluding pharmaceutical aides and unqualified auxiliary staff. For conciseness and international readability, this collective is uniformly referred to as “pharmacists” throughout this article.

### 2.2 Regional classification

China’s administrative division comprises 34 provincial-level units, including 23 provinces, 5 autonomous regions, 4 municipalities, and the Hong Kong and Macao Special Administrative Regions. Rural areas encompass county-level administrative units and their subordinate townships or villages, while urban areas refer to city-level administrative units and their corresponding urban districts.

For analytical purposes, mainland China is categorized into three broad regions—Eastern, Central, and Western—based on levels of per capita Gross Domestic Product (GDP).

The Eastern Region includes 11 provincial-level units: Beijing, Tianjin, Hebei, Liaoning, Shanghai, Jiangsu, Zhejiang, Fujian, Shandong, Guangdong, and Hainan.

The Central Region encompasses 8 provinces: Shanxi, Jilin, Heilongjiang, Anhui, Jiangxi, Henan, Hubei, and Hunan.

The Western Region consists of 12 provincial-level units: Chongqing, Sichuan, Guizhou, Yunnan, Tibet, Shaanxi, Gansu, Qinghai, Ningxia, Xinjiang, Guangxi, and Inner Mongolia.

### 2.3 Data resource

Data on pharmacists and healthcare institutions were sourced from the statistical yearbook series compiled by the National Health Commission of China. Specifically, we utilized data from the following yearbooks: the *National Health Statistics Yearbook (2008–2012)*, the *National Health and Family Planning Statistics Yearbook (2013–2017)*, and the *National Health Statistics Yearbook (2018–2024)*.

National and provincial demographic and economic data were obtained from the *Administrative Divisions Handbook of the People’s Republic of China* and *the China Statistical Yearbook (2007– 2024)*. It is important to note that, as these yearbooks typically publish prior-year data, figures for 2007 are contained within the 2008 edition, and this pattern applies correspondingly to other years.

Systematic, province-level data for pharmacists across all 31 provinces, autonomous regions, and municipalities directly under the central government of mainland China have only been available in the *China Health Statistics Yearbook since 2007*. While aggregate national totals for pharmacists can be traced to earlier years, this study uses 2007 as the unified starting point for all analyses involving inter-provincial comparisons to ensure temporal comparability across all analytical dimensions.Descriptions of national aggregate trends, however, incorporate data from earlier years where available. Furthermore, Hong Kong, Macao, and Taiwan were excluded from the analysis due to constraints in data availability and differences in statistical reporting formats.

### 2.4 Indicators

Quantity-related indicators included the total number of pharmacists and the number of pharmacists per 10,000 population, the latter calculated as the year-end total of pharmacists in healthcare institutions divided by the year-end resident population, multiplied by 10,000.

Quality-related indicators encompassed educational attainment (categorized as secondary specialized school or below, associate degree, bachelor’s degree, or master’s degree and above), professional title (senior, intermediate, or junior), and years of practice (<5, 5–9, 10–19, 20–29, or ≥30 years).

Structural indicators described the workforce composition, including gender distribution (male or female) and age distribution, where pharmacists were stratified into five groups: under 25 years, 25– 34 years, 35–44 years, 45–54 years, and 55 years or older.

Geographical distribution was assessed using the Gini coefficient, the Theil index, and hotspot analysis.

### 2.5 Data analysis

#### 2.5.1 Data Preprocessing

*The China Health Statistics Yearbook* underwent a revision in the classification criteria for healthcare institutions around 2009. To ensure comparability across the time series, data from before and after this change were harmonized. Specifically, the more granular institutional classifications for years prior to and including 2009 were recategorized according to the post-2009 framework. Consequently, data for all years were unified into four major categories: hospitals, primary healthcare institutions, professional public health institutions, and other healthcare institutions.

Furthermore, the definition of urban and rural areas in the *China Health Statistics Yearbook* was revised around 2010. The pre-2010 classification was based on an administrative hierarchy using “city” and “county” criteria, whereas the post-2010 classification adopted a geographical entity-based “urban” and “rural” criteria. As these two criteria are not directly comparable, analyses of urban-rural disparities in this study were conducted and discussed separately for two periods—2002–2009 and 2010–2023—to accurately reflect trends under their respective statistical standards.

Additionally, national-level structural data for pharmacists (including gender, age, education, and professional title) were missing for the years 2003, 2004, 2006, 2007, and 2008. To maintain the continuity of the time series, data for these missing years were imputed using the linear interpolation method.

#### 2.5.2 Equity and Spatial Analysis

The Gini coefficient is a statistical measure derived from the Lorenz curve, reflecting the degree of equity in social income distribution [19]. It was employed in this study to assess the equity of pharmacist resource allocation. The formula is as follows:

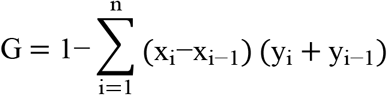

In this formula, G represents the Gini index, n represents the total number of regions, x_i_ represents the cumulative proportion of human resources in the i-th region, and y_i_ represents the cumulative proportion of CDC human resources in the i-th region. The value of the Gini coefficient ranges from 0 to 1. A coefficient approaching 0 indicates a more equitable distribution of pharmacist human resources among regions; conversely, a coefficient approaching 1 indicates a more concentrated allocation and poorer equity. It is generally accepted that a Gini coefficient below 0.2 represents a highly equitable distribution, a range of 0.2–0.3 indicates relative equity, 0.3–0.4 indicates relative inequity, and values exceeding 0.4 are considered highly inequitable [20-21].

To provide a deeper analysis of inequality, the Theil index was used to decompose the overall disparity into two primary components: inter-provincial disparity (T1L) and intra-provincial disparity (T2L). The formula is as follows:

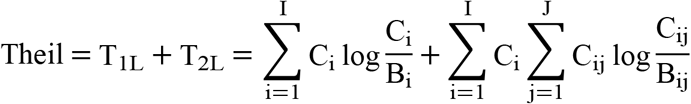

In this formula, B_i_ represents the total number of pharmacists in province i, and C_i_ represents the total resident population of province i. For measuring intra-provincial disparity, B_ij_ represents the proportion of pharmacists in urban or rural area j to the total number of pharmacists in province i, and C_ij_ represents the proportion of the population in urban or rural area j to the total population of province i. Furthermore, by calculating the contribution of each province’s Theil index to the total index, the degree to which intra-provincial disparities influence the overall imbalance can be determined. The value of the Theil index ranges from 0 to 1, with a lower value indicating a more equitable distribution of pharmacists [22].

Given that traditional econometric methods are insufficient to fully capture the spatial characteristics of pharmacist distribution, this study introduced spatial analysis techniques. Specifically, the global Moran’s I was employed to quantify the spatial heterogeneity and clustering of human resources, while hotspot analysis was used to visualize the distribution pattern, enabling more precise identification of regions with imbalanced resource allocation. The global Moran’s I evaluates the spatial autocorrelation of pharmacist distribution, with its statistical significance indicating the pattern of spatial association. The formula is as follows:

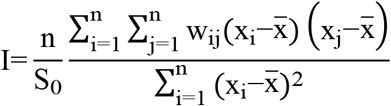

In the formula, n represents the total number of provinces; x_i_ and x_j_ represent the pharmacist density of province i and province j, respectively; 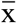 represents the mean of the dataset; w_ij_ represents the spatial weight value; and S_0_ represents the sum of all spatial weights, calculated as:

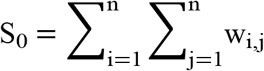

A statistically significant spatial autocorrelation is indicated when the p-value of the test is less than 0.05. This study calculated the global Moran’s I for pharmacist density across China’s 31 provinces from 2007 to 2023. The index ranges from −1 to 1, with its magnitude measuring the strength of spatial association. A value approaching +1 indicates strong positive spatial clustering, a value approaching −1 signifies significant spatial dispersion (or negative autocorrelation), and a value around 0 suggests that the distribution of pharmacists is spatially random [23].

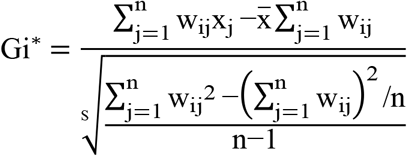

The calculated Gi* values were converted into standardized Z-scores. Provinces were then classified into three spatial clustering patterns based on the statistical significance assessed at the 95% confidence level (|Z| > 1.96): significant hotspots (Z>1.96, indicating high-value clustering), significant coldspots (Z<-1.96, indicating low-value clustering), and non-significant areas(|Z|≤1.96). A higher positive Z-score signifies that the value in a given area is significantly higher than the surrounding average, whereas a lower negative Z-score indicates it is significantly lower.

To enhance visualization, a graduated color scheme was applied in maps created using ArcGIS 10.8 (ESRI, CA, USA): dark red for high positive Z-scores (significant hotspots) and dark blue for low negative Z-scores (significant coldspots), intuitively illustrating patterns of resource agglomeration and dispersion. In constructing the spatial weight matrix, technical adjustments were made to account for the lack of land adjacency for Hainan Province. All calculations for the hotspot analysis were performed using the spdep package in R software (version 4.3.1).

### 2.6 Software tools

Descriptive statistical analyses of the workforce’s quantity, quality, and structural characteristics were conducted using Microsoft Excel 2017 (Microsoft, WA, USA). The Theil index and global Moran’s I were calculated with Stata 17 (StataCorp, TX, USA). Spatial visualization and autocorrelation analysis of pharmacist resources were carried out using ArcGIS 10.8 and GraphPad Prism (version 10.6). For all statistical tests, a P-value of less than 0.05 was defined as the threshold for statistical significance.

## 3 Methods

### 3.1 Quantity of Pharmacists

As shown in Figure 1, the total number of pharmacists in healthcare institutions in mainland China demonstrated a growth trend from 2002 to 2023, increasing from 357,700 to 569,500, with an average annual growth rate of 2.2%. A disaggregated view shows that the number of pharmacists in urban areas rose from 146,000 to 325,600, while the number in rural areas increased from 61,600 to 243,900. Consequently, the density of pharmacists per 10,000 population grew from 2.78 in 2002 to 4.04 in 2023. This increase was observed in both urban settings, where the density rose from 1.14 to 2.31 per 10,000 population, and rural settings, where it increased from 0.48 to 1.73 per 10,000 population.

**Figure 1.**
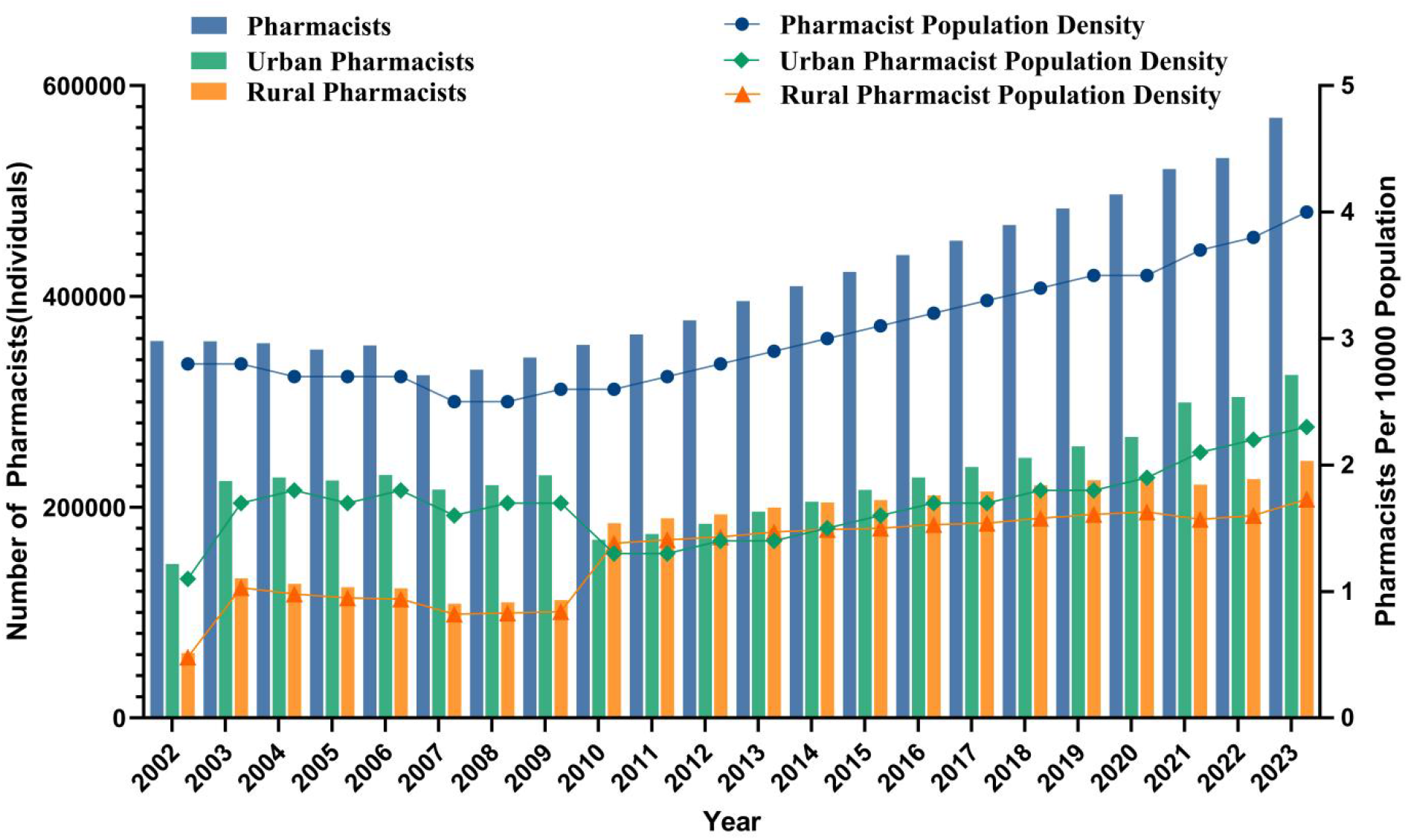
Trends in the number of Pharmacists in Mainland China from 2002 to 2023

A notable decline in the total number of institutional pharmacists in 2007 is likely attributable to a revision in the statistical criteria used by the health authorities that year, whereby pharmacy assistants working in hospitals without professional titles were excluded from the count. Furthermore, the apparent sharp decrease in urban pharmacists alongside a rapid increase in rural pharmacists in 2010 can be explained by a major change in the urban-rural classification standard adopted by the China Health Statistics Yearbook around that time. The pre-2010 classification was based on the administrative hierarchy of “city” and “county,” whereas the post-2010 classification shifted to a geographical entity-based definition of “urban” and “rural” areas.

As illustrated in Figure 2, from 2002 to 2023, the numbers and population densities (per 10,000 population) of physicians, nurses, and pharmacists in China all exhibited growth trends, albeit with markedly divergent growth rates. The physician workforce increased from 1.844 million to 4.7821 million, corresponding to an average annual growth rate of 4.6%. The nursing workforce expanded from 1.2465 million to 5.6371 million, with an average annual growth rate of 7.4%. In contrast, the pharmacist workforce grew from 357,700 to 569,500, but its growth rate (2.2% annually) was substantially lower than that of both physicians (4.6%) and nurses (7.4%).

**Figure 2.**
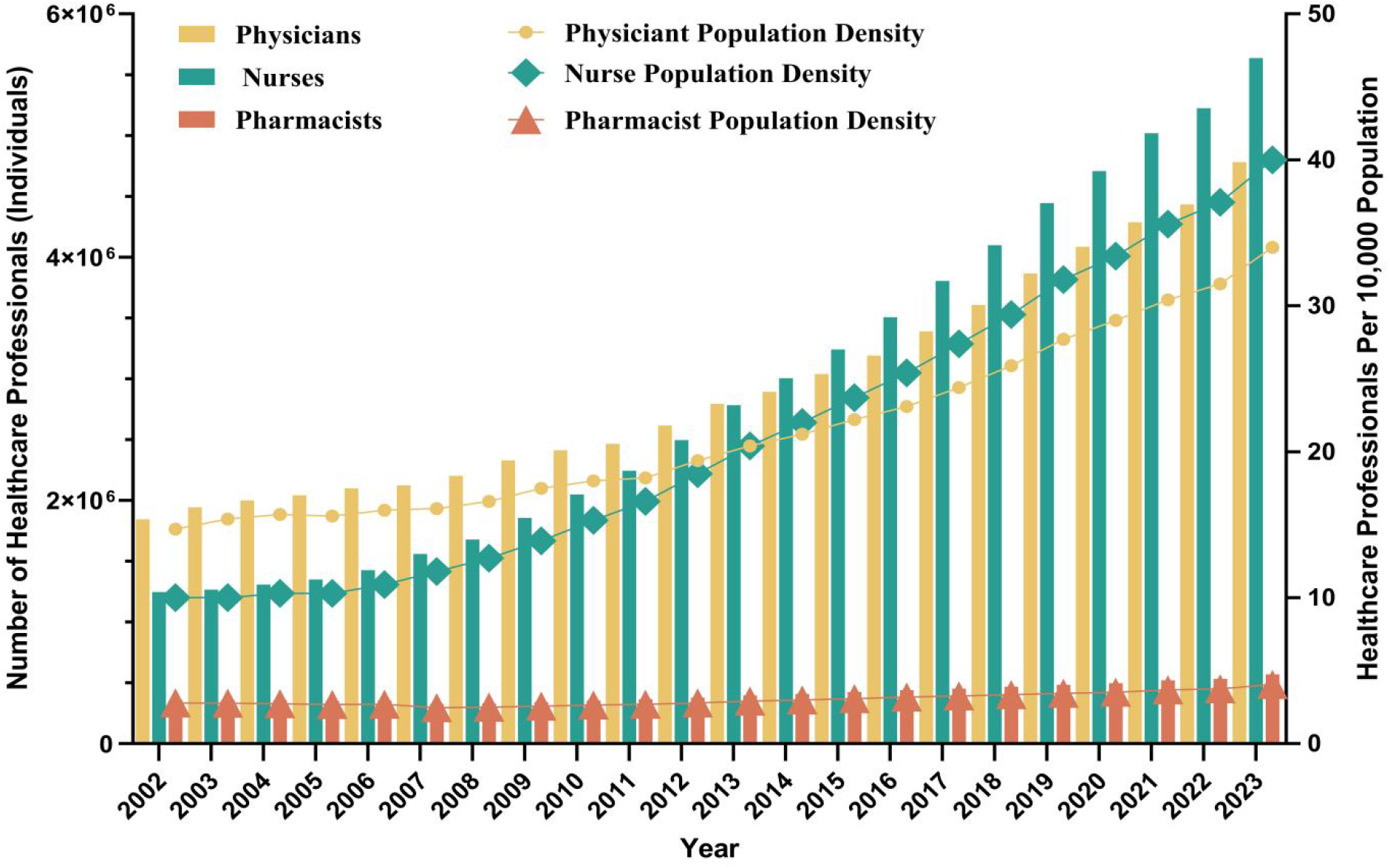
Comparative Trends in the Number and Density of Physicians, Nurses, and Pharmacists in China, 2002–2023

A similar pattern was observed for workforce density. From 2002 to 2023, physician density increased from 14.7 to 34 per 10,000 population, nurse density rose from 10 to 40 per 10,000 population, and pharmacist density grew from 2.78 to 4.04 per 10,000 population. A critical finding is the notable decline in the relative proportions of pharmacists to other health professionals. The pharmacist-to-physician ratio decreased from 1:5.15 to 1:8.39, and the pharmacist-to-nurse ratio declined from 1:3.48 to 1:9.90. These data indicate that during the overall expansion of China’s health human resources over the past two decades, the growth of the pharmacist workforce has lagged, resulting in a significant relative contraction of its size within the healthcare team.

### 3.2 Structure of the Pharmacist Workforce

As presented in Table 1, in 2002, male pharmacists accounted for 40.3% of the total, compared to 59.7% for females. By 2023, the gender distribution had shifted further towards females, with the proportion of female pharmacists rising to 70.8%. Regarding age structure in 2023, the proportion of young pharmacists under 25 years old decreased compared to 2002 (4.0% vs. 11.3%), whereas the proportion of pharmacists aged 55 and above increased significantly (8.9% vs. 2.4%). Concurrently, pharmacists aged 25–34 constituted the largest proportion in 2023 (34.5%), and the share of those aged 35–44 remained relatively stable (31.4% in 2023 vs. 32.4% in 2002).

**Table 1.**
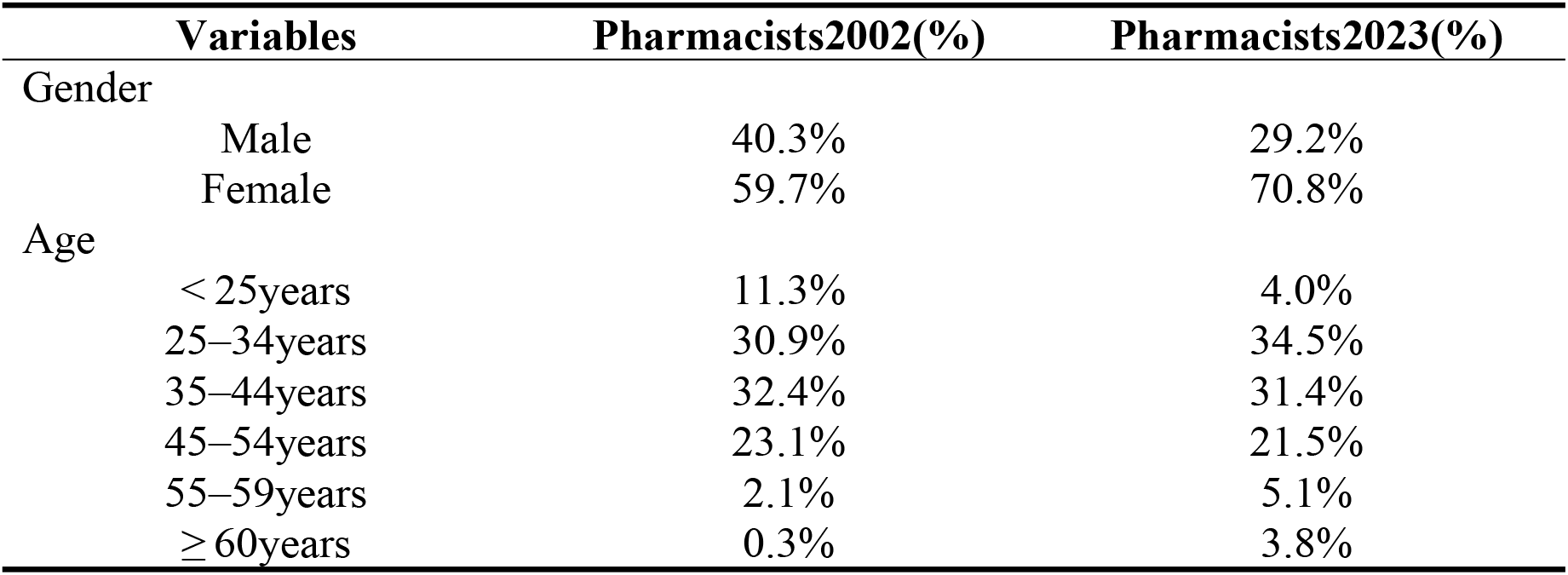
Gender and Age Distribution of the Pharmacist Workforce in Chinese Healthcare Institutions, 2002 and 2023.

### 3.3 Quality of the Pharmacist Workforce

As detailed in Table 2, the educational attainment of the pharmacist workforce improved markedly over the two decades. In 2002, only 0.2% held a master’s degree or higher, while 99.8% had a bachelor’s degree or lower (including associate degrees). By 2023, the proportion of pharmacists with at least a bachelor’s degree had risen to 48.9%, of which 6.1% held a master’s degree or higher.

**Table 2.** Educational Attainment, Professional Titles, and Years of Practice of the Pharmacist Workforce in Chinese Healthcare Institutions, 2002 and 2023.

| Variables | Pharmacists2002(%) | Pharmacists2023(%) |
| --- | --- | --- |
| Education levels |  |  |
| Master or above | 0.2% | 6.1% |
| Baccalaureate | 5.2% | 42.8% |
| Associate degree or below | 94.6% | 51.1% |
| Professional titles |  |  |
| Advanced titles | 2.4% | 6.6% |
| Intermediate titles | 20.1% | 24.6% |
| Preliminary titles | 69.2% | 36.9% |
| Others | 8.3% | 2.8% |
| Work experiences |  |  |
| < 5years | 13.2% | 18.4% |
| 5–9years | 15.0% | 19.8% |
| 10–19years | 27.8% | 29.8% |
| 20–29years | 31.5% | 17.0% |
| ≥ 30years | 12.1% | 15.0% |

Regarding professional titles, 6.6% of pharmacists held senior titles in 2023, 24.6% held intermediate titles, and 36.9% held junior titles. Compared to 2002, the shares of both senior (2.4%→ 6.6%) and intermediate (20.1% → 24.6%) titles increased, whereas the proportion of junior titles decreased substantially (69.2%→ 36.9%).

In terms of work experience, pharmacists with fewer than 10 years of practice accounted for 38.2% of the workforce in 2023 (compared to 28.2% in 2002). Conversely, those with 30 or more years of experience constituted 15.0% (compared to 12.1% in 2002).

### 3.4 Distribution of Pharmacists

As shown in Figure 3, the Gini coefficient for pharmacist resource allocation across China’s 31 provincial-level regions exhibited a consistent downward trend from 2007 to 2023, decreasing gradually from 0.1448 to 0.0928. Although a minor rebound occurred in 2011 (0.1277), the overall fluctuation was limited, and the declining trajectory remained pronounced. Notably, after 2015, the Gini coefficient stabilized below 0.11 and reached its lowest value of 0.0928 in 2023, indicating that the equity of pharmacist distribution at the provincial level has progressively improved.

**Figure 3.**
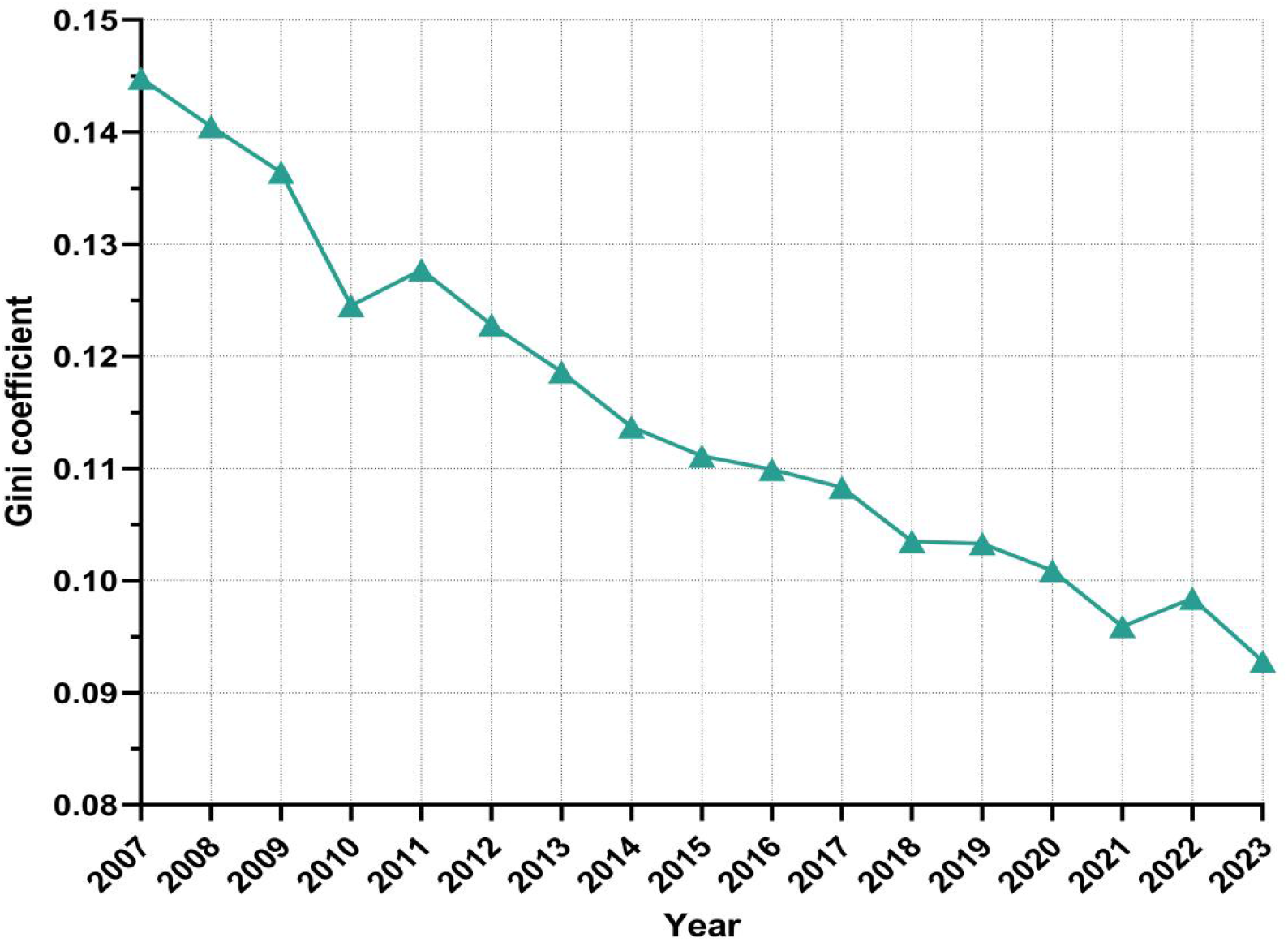
Trend in the Gini Coefficient of Pharmacist Resource Allocation in China (2007–2023) This line graph illustrates the dynamic change in the Gini coefficient for pharmacist resource allocation across China’s 31 provincial-level regions from 2007 to 2023. The coefficient shows a consistent decline from 0.1448 in 2007 to 0.0928 in 2023, despite a slight increase in 2011, indicating a steady improvement in the equity of pharmacist distribution over the study period. (Data source: China Health Statistics Yearbook)

The temporal trend of the Theil index for pharmacist resources in Chinese healthcare institutions from 2007 to 2023 was calculated (Figure 4). The evolution of the total Theil index (Theil L) can be broadly delineated into three phases. During 2007–2011, the index decreased rapidly from 0.116 to approximately 0.05. A pronounced anomalous peak appeared in 2012 (0.255). From 2013 onward, the index receded and fluctuated within a relatively low range of 0.05–0.066.

**Figure 4.**
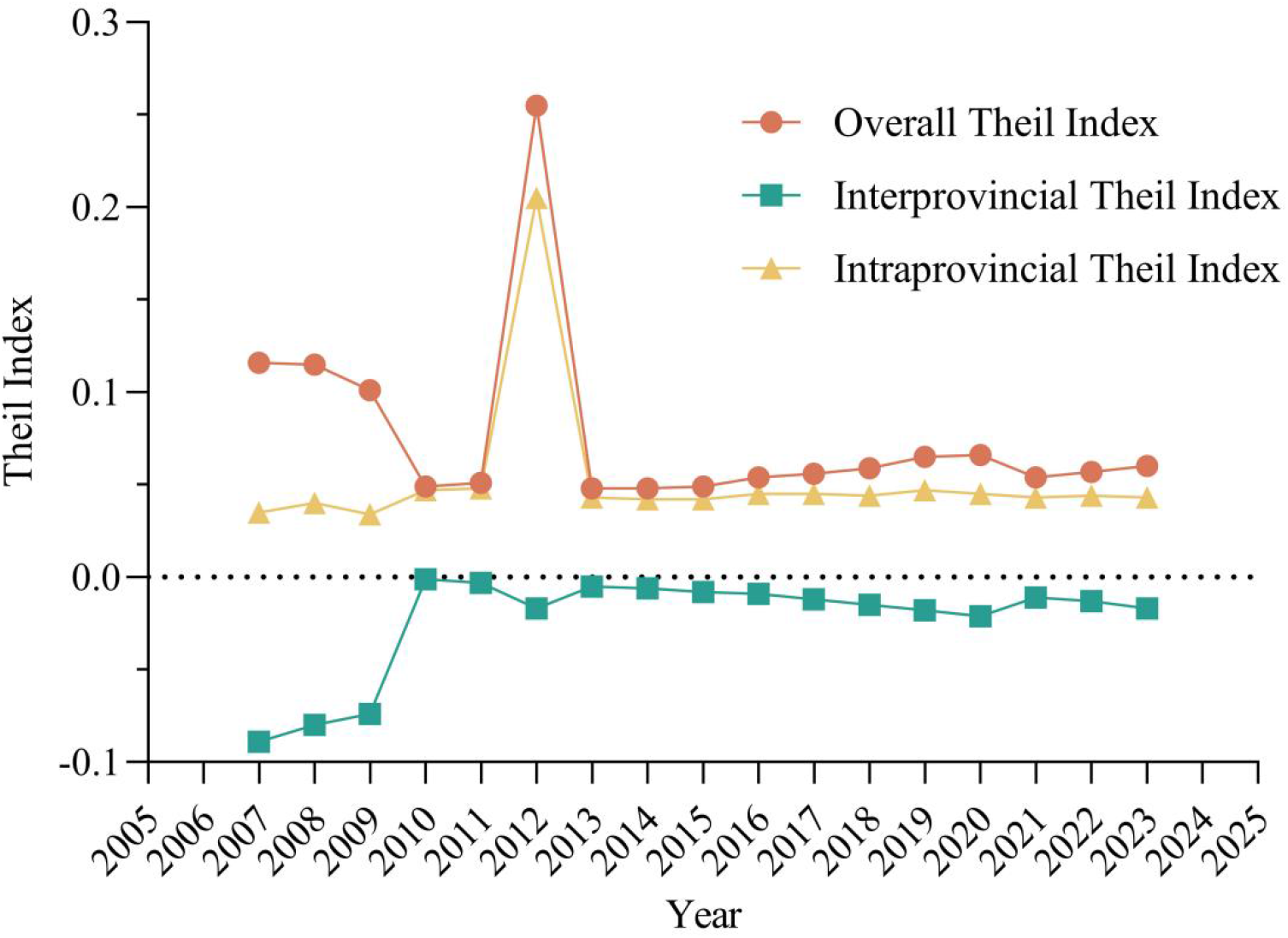
Trends in the Theil Index and Its Decomposition for Pharmacist Resource Allocation in Chinese Healthcare Institutions, 2007–2023. This figure illustrates the trends of the total Theil index (Theil L), the between-group disparity (T_1L_, urban-rural), and the within-group disparity (T_2L_, within urban and rural areas) for pharmacist resource allocation in China from 2007 to 2023. The between-group disparity (T_1L_) remained negative throughout the study period, located below the zero reference line (y=0). Data were sourced from the *China Health Statistics Yearbook (2008–2024)*.

Decomposition of the index reveals that the between-group component (T_1L_), representing urban-rural disparity, was negative throughout the study period(Table 3). This indicates that the pharmacist density per 10,000 population was higher in rural areas than in urban areas. Conversely, the within-group component (T_2L_), representing inter-provincial disparities within urban and rural areas, remained positive and constituted the primary source of the overall inequality.

**Table 3.** Theil Index and Its Decomposition for Pharmacist Resource Allocation in Chinese Healthcare Institutions, 2007–2023.

| Year | Theil L | T1L<br>(Between) | T1L/TL<br>(%) | T2L<br>(Within) | T2L/TL<br>(%) |
| --- | --- | --- | --- | --- | --- |
| 2007 | 0.116 | -0.089 | -76.80 | 0.035 | 29.654 |
| 2008 | 0.115 | -0.080 | -69.278 | 0.040 | 34.909 |
| 2009 | 0.101 | -0.074 | -73.110 | 0.034 | 33.382 |
| 2010 | 0.049 | -0.001 | -2.637 | 0.047 | 96.495 |
| 2011 | 0.051 | -0.003 | -5.686 | 0.048 | 93.482 |
| 2012 | 0.255 | -0.017 | -6.720 | 0.205 | 80.631 |
| 2013 | 0.048 | -0.005 | -10.023 | 0.043 | 89.488 |
| 2014 | 0.048 | -0.006 | -12.913 | 0.042 | 86.919 |
| 2015 | 0.049 | -0.008 | -15.317 | 0.042 | 84.791 |
| 2016 | 0.054 | -0.009 | -17.352 | 0.045 | 82.341 |
| 2017 | 0.056 | -0.012 | -20.683 | 0.045 | 79.651 |
| 2018 | 0.059 | -0.015 | -25.948 | 0.044 | 74.708 |
| 2019 | 0.065 | -0.018 | -27.509 | 0.047 | 72.937 |
| 2020 | 0.066 | -0.021 | -31.816 | 0.045 | 68.453 |
| 2021 | 0.054 | -0.011 | -19.997 | 0.043 | 79.500 |
| 2022 | 0.057 | -0.013 | -22.343 | 0.044 | 77.416 |
| 2023 | 0.060 | -0.017 | -27.799 | 0.043 | 71.758 |

Note: The total Theil index (Theil L) can be decomposed into between-group disparity ( $T_{1L}$ , between urban and rural areas) and within-group disparity ( $T_{2L}$ , within urban and within rural areas, respectively). A negative value for $T_{1L}$ indicates that the ratio of pharmacist density in rural areas to the national average is higher than that in urban areas. The contribution rate represents the percentage of the absolute value of each component's disparity relative to the total index. The notable surge in the total index in 2012 is likely associated with the adjustment in statistical criteria for that year. Data were processed and calculated by the authors; the raw data were obtained from the *China Health Statistics Yearbook (2008–2024)*.

The anomalous peak in 2012 is likely attributable to the significant revision of the urban-rural classification criteria in the *China Health Statistics Yearbook* for that year. The subsequent stabilization of the index trend suggests that the equity of resource allocation has been in a state of continuous improvement since 2013.

This study employed the global Moran’s I to assess the spatial autocorrelation characteristics of pharmacist human resource density across 31 provinces in mainland China from 2007 to 2023. The results indicate that the spatial distribution pattern of pharmacist resources changed over time (Figure 5). Specifically, the trend in the spatial distribution of pharmacist density can be divided into two distinct phases. In the first phase (2007–2011), the global Moran’s I showed a gradual decline,decreasing from a peak of 0.254 in 2007 to 0.151 in 2011. During this period, pharmacist density exhibited a statistically significant positive spatial autocorrelation (\**P*< 0.05), indicating a clustered distribution pattern and suggesting room for improvement in the spatial equity of resource allocation.

**Figure 5.**
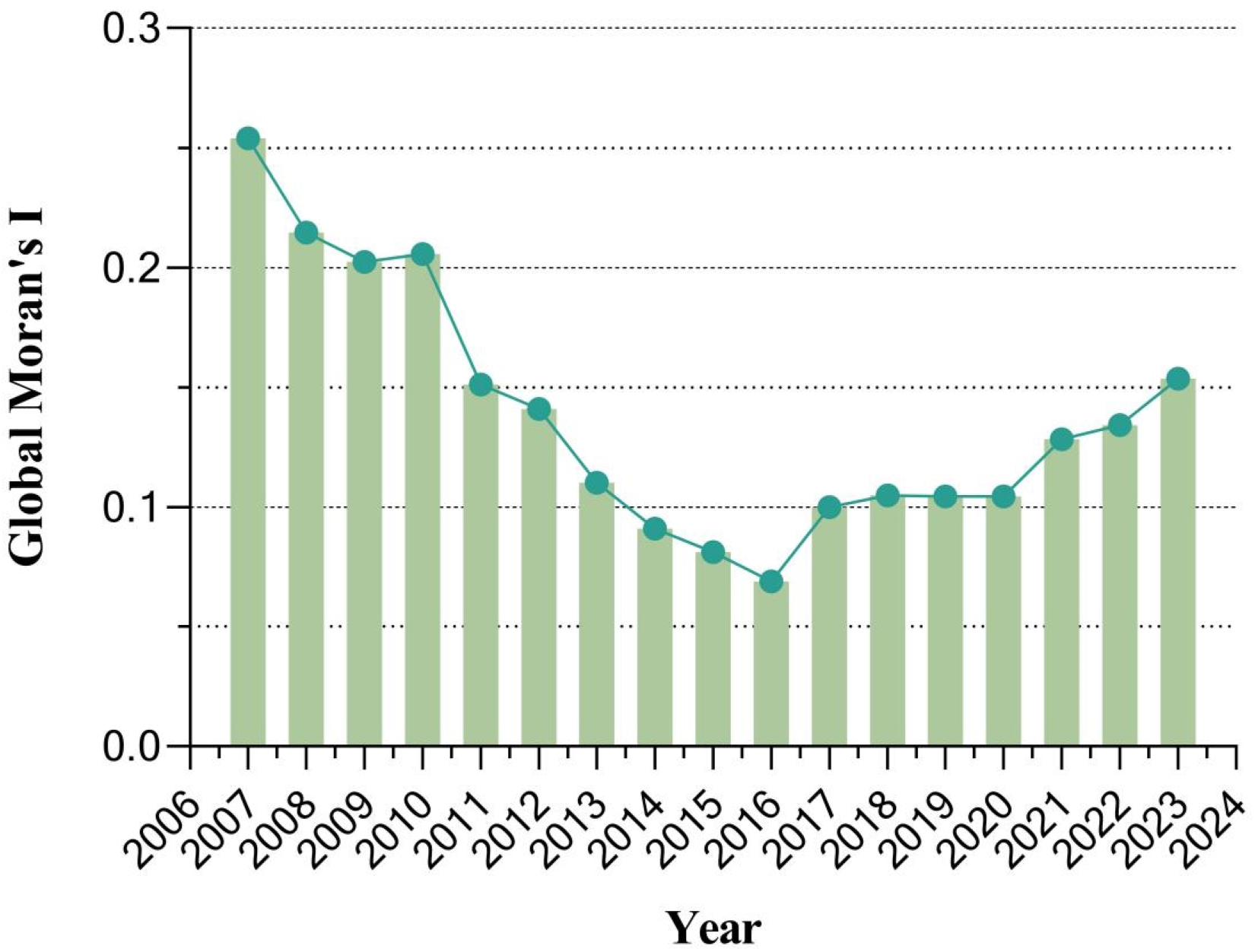
Declining Trend in Spatial Autocorrelation of Pharmacist Density Across Chinese Provinces, 2007-2023

In the second phase (2012–2023), the global Moran’s I fluctuated between 0.069 and 0.154, with pharmacist density showing no significant spatial autocorrelation (*P* > 0.05). This suggests that the spatial distribution of pharmacists tended toward randomization during this later period, reflecting an improvement in distributional equity. This observed shift may be associated with recent policy orientations aimed at strengthening pharmaceutical care at the primary level and promoting the decentralization of pharmacist resources.

Table 4 presents the annual calculation results of the global Moran’s I from 2007 to 2023. In 2007, pharmacist resources exhibited the strongest positive spatial autocorrelation (Moran’s I = 0.2539, \*\**P* < 0.01). Subsequently, the strength of spatial autocorrelation diminished annually, remaining statistically significant until 2011 (Moran’s I = 0.1512, \**P* < 0.05). For all years from 2012 onward, the P-values exceeded 0.05, indicating that the spatial distribution of pharmacist resources in these years did not show a statistically significant pattern of either clustering or dispersion. All calculations were based on a spatial weight matrix constructed using the “Queen” contiguity definition (row-standardized), with technical adjustments applied to account for the lack of land adjacency for Hainan Province.

**Table 4.**
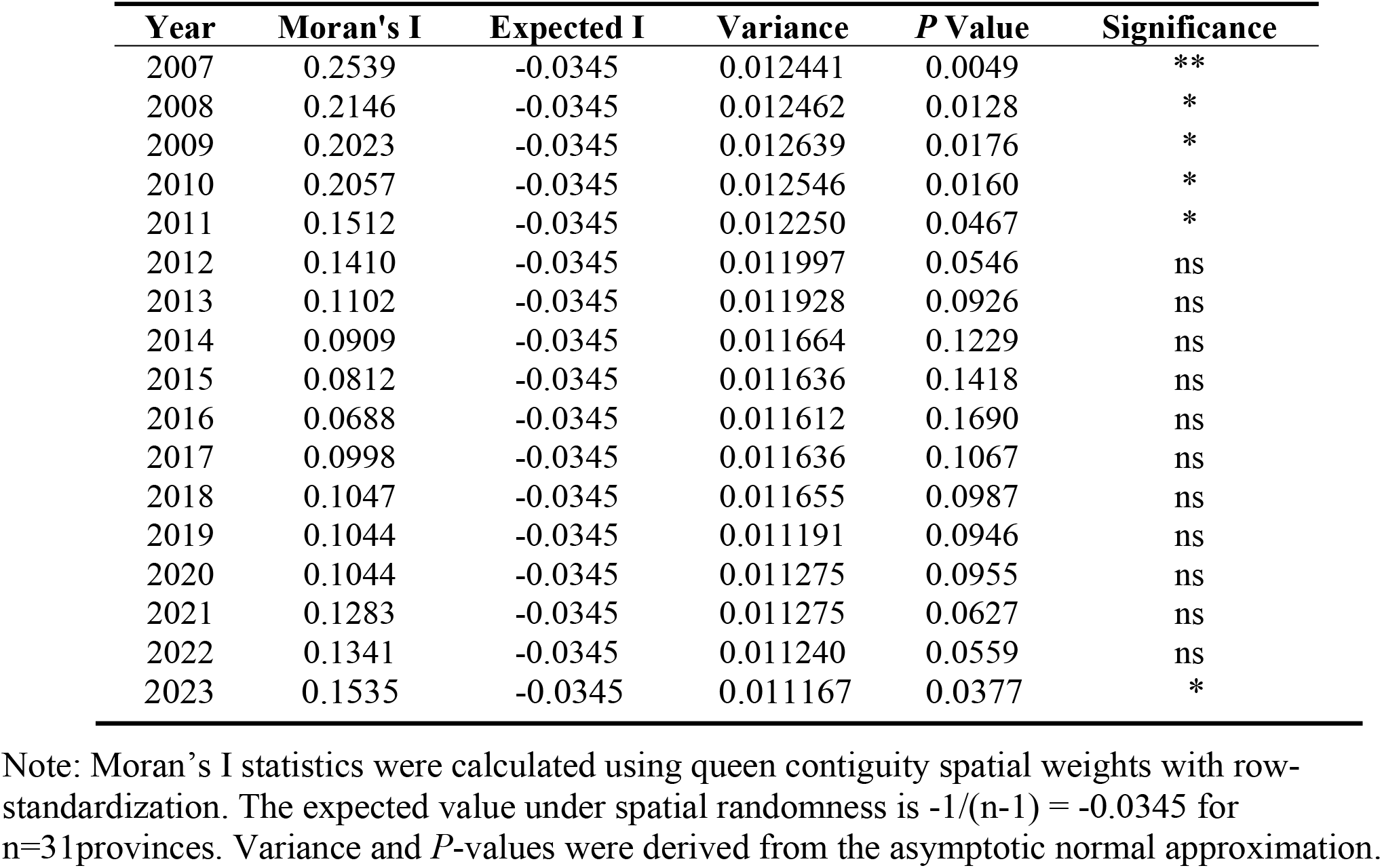
Global Moran’s I statistics for pharmacist density across 31 provinces in mainland China, 2007-2023.

The results of the Getis-Ord Gi hotspot analysis (Figure 6) reveal a dynamic evolution in the intensity of spatial agglomeration of pharmacist resources in China from 2007 to 2023, characterized by a transition from “strong clustering” to “weak clustering” and finally to “low clustering.”

**Figure 6.**
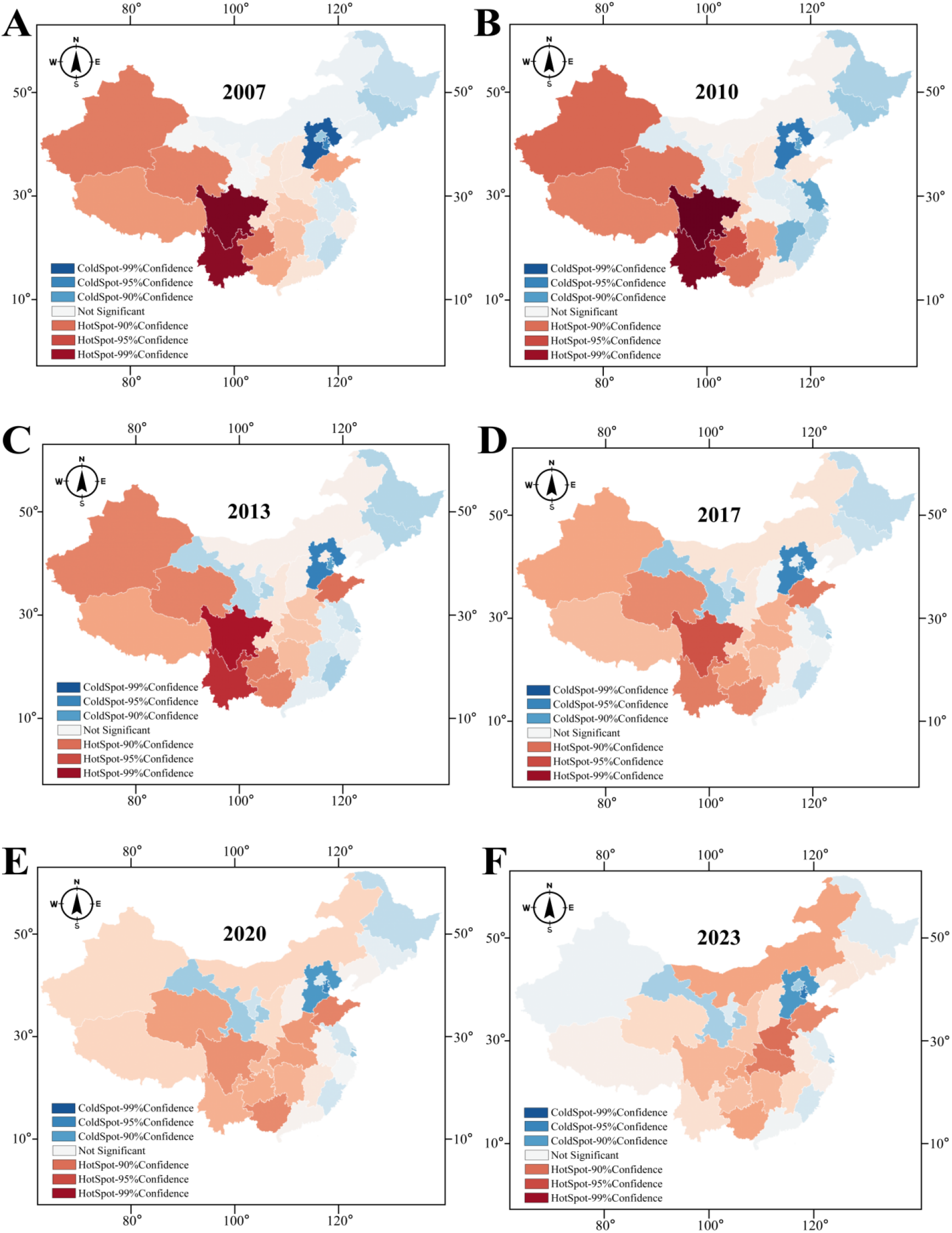
Spatiotemporal evolution of Getis-Ord Gi* hotspot analysis for pharmacist human resource density in China, 2007-2023. Panels A–F correspond to the distribution results for the years 2007, 2010, 2013, 2017, 2020, and 2023, respectively. The color gradient reflects the continuous variation in Z-scores: blue hues (Z-score < 0) represent coldspots (low-density clusters), and red hues (Z-score > 0) represent hotspots (high-density clusters). Statistical significance is determined at the 95% confidence level (|Z| > 1.96, *P* < 0.05): dark colors (dark blue/dark red) indicate areas of clustering at the 99% confidence level, while light colors (light blue/light red) indicate clustering at the 95% confidence level. Areas labeled “Not Significant” represent regions with no statistically significant spatial clustering. The spatial weight matrix was constructed based on the rule of geographical contiguity and was standardized.

During the period 2007–2010, spatial clustering was most pronounced. Well-defined core areas of strong hotspots with 99% confidence (depicted in dark red) were evident. Simultaneously, extensive coldspot clusters with 99% confidence (dark blue) were observed in northern regions. The majority of other areas were identified as 95% confidence hotspots (in shades of red), while areas exhibiting no significant clustering accounted for a minimal proportion of the total.

From 2013 to 2017, the intensity of spatial agglomeration progressively weakened. The spatial extent of the high-confidence (99%) hotspot regions began to contract, with some peripheral areas downgrading to lower-confidence (95%) hotspots. Concurrently, the previously extensive coldspot clusters in the north became more fragmented, giving way to scattered, isolated coldspots at the 95% confidence level. The areal proportion of regions with no significant clustering increased during this phase.

By the period 2020–2023, high-confidence clustering had largely dissipated. The spatial aggregation of coldspots further dispersed, leaving only a few isolated, weak coldspot areas at the 95% confidence level. Most of the country displayed no statistically significant spatial clustering, indicating that the spatial agglomeration intensity of pharmacist density in healthcare institutions had substantially diminished.

## 4 Discussion

### 4.1 Analysis of Pharmacist Quantity

This study reveals a significant structural shortage in the pharmacist human resources within Chinese healthcare institutions. From 2002 to 2023, despite the increase in the total number of pharmacists, the density remained at only 4.04 pharmacists per 10,000 population. This figure is substantially lower than both the global average (9.6 per 10,000 population) and the level observed in high-income countries (12.1 per 10,000) [24]. To reach the global average, an additional approximately 770,000 pharmacists would be required.

The scale of the pharmaceutical market underscores the magnitude of their responsibility. Data indicate that in 2024, the total value of drug sales in China was approximately 18,638 billion yuan. Sales in healthcare institutions accounted for 11,152 billion yuan, and sales in public primary healthcare institutions accounted for 1,747 billion yuan, together constituting about 69.2% of the total drug sales. In contrast, sales in retail pharmacies accounted for about 30.8% [25]. Notably, the number of pharmacists in healthcare institutions is 175,000 fewer than the number of licensed pharmacists in retail pharmacies [26]. This smaller workforce, however, is tasked with ensuring the supply and promoting the rational use of nearly 70% of the nation’s drugs (predominantly prescription medications) [27].

Due to this severe understaffing, the majority of institutional pharmacists are compelled to focus their efforts on managing high-intensity, repetitive drug supply and dispensing tasks. Consequently, they have insufficient time and energy to dedicate to clinical pharmacy services. This limitation likely hinders their full potential in advancing key healthcare system goals such as the implementation of tiered diagnosis and treatment, effective medical insurance cost containment, and the overall enhancement of rational drug use.

In 2018, China issued the *Regulations on Pharmaceutical Affairs Management in Medical Institutions*, which stipulate that the proportion of pharmaceutical professionals among all healthcare professionals within an institution should not be less than 8% to ensure rational medication use and patient safety [5]. This study found that during the overall expansion of China’s health human resources over the past two decades, the growth of the pharmacist workforce has lagged. Its average annual growth rate (2.2%) was considerably lower than that of physicians (4.6%) and nurses (7.4%). Consequently, the deployment of pharmacists in healthcare institutions has consistently fallen short of this standard, with their proportion declining yearly and the gap to the 8% target widening continuously.

This lag has resulted in a sustained contraction of their relative size within the healthcare team. The ratio of pharmacists to physicians in healthcare institutions is substantially lower than that in developed countries (typically ranging from 1:2 to 1:4) and even below the levels in many middle-income countries (typically 1:4 to 1:5) [28]. This comparison reveals that the shortage of pharmacist human resources in China is not merely an issue of absolute numbers but, more critically, a structural disproportionality within the healthcare team [29-30]. Such an imbalance may render pharmacists unable to provide adequate professional and technical pharmaceutical support to physicians and nurses in the face of rapidly increasing clinical service volumes, which could potentially compromise inter-professional collaboration efficiency and patient medication safety. Future analyses should further investigate the underlying causes of this pronounced international disparity.

### 4.2 Analysis of Pharmacist Structure

This study indicates that the gender composition of China’s pharmacist workforce in healthcare institutions has continued to shift towards a female majority, with the proportion of female pharmacists rising significantly from 59.7% in 2002 to 70.8% in 2023. This trend aligns with broader global shifts in the gender composition of health professional fields. The rising proportion of females in health professions in China may be attributed to enhanced access to and equity in higher education for women, alongside the profession’s emphasis on communication and meticulousness, which may appeal to female career preferences.

Concurrently, the age distribution reveals a declining proportion of younger practitioners. This phenomenon is directly linked to a decrease in the profession’s attractiveness. Several systemic factors contribute to this: first, operational cost controls in hospitals (driven by policies like the zero-markup drug policy and DRG/DIP payment reforms) and constraints on authorized staffing levels have led many institutions to prioritize hiring for “frontline” positions like physicians and nurses, often freezing or reducing pharmacist recruitment. Second, limited opportunities for professional advancement often confine pharmacists to basic drug supply roles, with few chances to engage in clinical decision-making or manage complex medication therapies. Furthermore, core pharmacist services that demonstrate professional value, such as prescription review and medication education, often lack dedicated fee items and carry insufficient weight in performance evaluations. This disconnect between effort, value recognition, and compensation dampens the morale of current pharmacists and significantly reduces the field’s appeal to new graduates.

### 4.3 Quality of Pharmacists

This study identified a pronounced structural imbalance in the educational attainment and professional title composition of the pharmacist workforce in healthcare institutions. Specifically, individuals holding a bachelor’s degree or lower accounted for 93.9%, while only 6.1% possessed a postgraduate degree or above. In terms of professional titles, those with junior titles constituted 67.8% of the workforce, whereas a mere 7.0% held senior titles. This suboptimal structure constrains the capacity of pharmacists to deliver high-level clinical pharmaceutical care and to engage in scientific research and innovation.

### 4.4 Distribution of Pharmacists

This study employed an integrated approach, utilizing the Gini coefficient, Theil index, and spatial analysis methods to assess the equity and its evolution in the allocation of pharmacist resources within Chinese healthcare institutions from 2007 to 2023. The results demonstrate a continuous improvement in the equity of provincial-level allocation, a significant weakening of spatial agglomeration, and a higher relative density of pharmacists in rural areas compared to urban areas.

These observed changes in resource allocation exhibit a temporal correspondence with the rollout of relevant national policies during the same period. From 2007 to 2011, the Gini coefficient began to decline; however, the global Moran’s I remained statistically significant, indicating that a spatially clustered distribution of pharmacists persisted. This phase of initial improvement broadly coincided with the enhanced emphasis on pharmaceutical services within the healthcare system following the launch of the new healthcare reform in 2009, and more specifically, with the implementation of the *Regulations on Pharmaceutical Affairs Management in Medical Institutions* in 2011 [3-4].

In 2012, the Theil index exhibited an anomalous peak, and concurrently, the global Moran’s I lost its statistical significance from this year onward. This shift is primarily attributable to the revision of the urban-rural classification criteria in the statistical yearbooks for that year, which introduced a short-term disruption to data continuity.

Following 2013, all indicators demonstrated a more stable trend toward optimization. The Gini coefficient stabilized at a low level, and the spatial extent of high-confidence agglomeration areas in the hotspot analysis maps continued to diminish. These trends closely followed, in a temporal sequence, the issuance of key national policies such as the *Opinions on Accelerating the High-Quality Development of Pharmaceutical Care* (2018) and the *Opinions on Strengthening Pharmaceutical Affairs Management in Healthcare Institutions to Promote Rational Drug Use* (2020) [5-6]. These policies explicitly emphasize the development of pharmaceutical care at the primary level and the strengthening of the pharmacist workforce.

The findings of this study align with these policy imperatives. The observed higher relative density of pharmacists in rural areas compared to urban settings may be a tangible reflection of the policy-driven inclination of pharmaceutical service resources towards the grassroots level. Similarly, the weakening of the spatial agglomeration of pharmacist resources nationwide resonates with the overarching policy goal of achieving a more equitable geographical allocation of resources.

The within-group disparity (T_2L_) indicates that resource inequality among provinces within urban areas and, separately, among provinces within rural areas constitutes the predominant source of current inequality. Policy efforts should therefore prioritize narrowing the gaps in pharmacist resources both between provinces of differing economic levels and among different areas within individual provinces. Concurrently, policies must foster the synchronous enhancement of the professional competencies of the pharmacist workforce. These coordinated actions are essential to achieve tangible improvements in both the accessibility and quality of pharmaceutical care.

The phenomenon of higher relative pharmacist density in rural areas identified in this study is likely contextualized within the national policy background of continuously strengthening the primary health workforce. To improve healthcare accessibility in rural areas, national plans have guided resources toward the grassroots level and implemented multiple talent support initiatives, including the targeted tuition-free medical student program for rural areas [46-47]. Under this overarching policy orientation, the allocation of pharmacists—a crucial component of the primary health team— has also shown a trend of reinforcement in rural areas. This shift aligns with the concurrent general increase in the number of healthcare personnel in rural regions [7].

Future attention should focus on the professional competency structure, service models, and workload of the rural pharmacist workforce. This focus is necessary to evaluate the practical effectiveness of resource decentralization and to explore how subsequent policy support can translate the relative quantitative advantage into a tangible enhancement in the quality of pharmaceutical care.

## 5 Conclusion

This study presents a systematic analysis of the long-term evolution of the pharmacist workforce within healthcare institutions in mainland China. In terms of quantity, while the total number of pharmacists has sustained growth, its growth rate has been significantly lower than that of physicians and nurses. This disparity has led to a continuous contraction of their relative scale within the healthcare team, highlighting a pronounced issue of structural shortage. The workforce structure exhibits trends toward feminization and aging, with insufficient replenishment by younger professionals. Although the overall levels of educational attainment and professional titles have improved, the proportion of high-level talents remains low, potentially constraining the depth of development in clinical pharmaceutical care capabilities. Regarding geographical distribution, equity in allocation at the provincial level has shown continuous improvement, and spatial agglomeration has weakened, reflecting the positive effects of national macro-level resource allocation policies. However, disparities within urban and rural areas and between provinces remain the primary sources of current equity challenges.

Several limitations of this study should be noted. First, our analysis of pharmacist distribution is limited to the provincial level; future research incorporating municipal and county-level data would allow for a more granular and comprehensive analysis. Second, the structural data for pharmacists (e.g., gender, age) for certain years are imputed values, which may have introduced some bias and affected the precision of the longitudinal trend analysis. Third, this study focuses exclusively on pharmacists practicing within healthcare institutions and does not include pharmacists in retail pharmacy settings. Therefore, the findings may not fully represent the entire landscape of China’s pharmacist workforce. Finally, while this study observes a temporal association between improvements in resource allocation and major policy cycles, the observational design precludes the establishment of strict causality. More rigorous study designs are needed in the future to verify the net effects of specific policies.

## 7 Author Contributions

**Yuxiang Xia:** Conceptualization; methodology; investigation; data curation; formal analysis; writing– original draft; writing – review & editing; visualization; project administration; final approval of the version to be published; agreement to be accountable for all aspects of the work. **Lulu Sun:** Conceptualization; methodology; validation; formal analysis; visualization; writing – review & editing; final approval of the version to be published; agreement to be accountable for all aspects of the work. **Yingbo Zhao:** Conceptualization; methodology; supervision; writing – review & editing; final approval of the version to be published; agreement to be accountable for all aspects of the work.

## 8 Data availability statement

No data was generated by this study. The following existing data sources were used:· The China Health Statistics Yearbook (2008–2024) is available from the Statistical Information Center of the National Health Commission of China: https://www.nhc.gov.cn/mohwsbwstjxxzx/tjtjnj/list.shtml. Copies of the yearbooks can also be accessed via www.tjcn.org.The China Statistical Yearbook (2007–2024) is available from the National Bureau of Statistics of China: https://www.stats.gov.cn. Copies of the yearbooks can also be accessed via www.tjcn.org.

## 9 Acknowledgments

Not applicable.

